# Developing a Specialized Dravet Syndrome Ontology for Rare Disease Informatics and AI Applications

**DOI:** 10.64898/2026.07.01.26357055

**Authors:** Pedram Golnari, Katrina Prantzalos, Dipak P. Upadhyaya, Jeffrey Buchhalter, Satya S. Sahoo

## Abstract

Dravet syndrome (DS) is a severe developmental and epileptic encephalopathy whose clinical and research representation requires integration of heterogeneous knowledge spanning seizures, development, behavior, SUDEP/autonomic risk, genetics, comorbidities, electrophysiology, pharmacology, and drug responsiveness. We report the development of a DS-focused ontology created by expert-guided specialization of a previously published epilepsy ontology. Scope expansion was defined through a scientific advisory board, structured review meetings, and iterative ontology curation in OWL. The resulting resource reorganized DS content across nine major domains and expanded the publicly released ontology from the pre-extension baseline to the current BioPortal version. Beyond structural growth, the ontology was assessed through expert-guided curation and downstream task-based reuse, including two published ontology-enabled LLM studies and an ongoing ontology-derived DS knowledge graph and AI assistant platform. These results suggest that disease-focused ontology specialization can provide durable infrastructure for DS data harmonization, knowledge representation, and AI-enabled translational informatics.

## Introduction

Dravet syndrome is a rare developmental and epileptic encephalopathy that usually begins in infancy, is commonly associated with pathogenic variants in SCN1A,1-4 and evolves beyond seizures to include developmental, behavioral, autonomic, and treatment-related complications.^1, 2, 5-8^ Clinical management therefore depends on integrating heterogeneous information spanning seizure semiology, neurodevelopment, comorbidities, genetics, electrophysiology, pharmacology, and risk of premature death.^1, 8^ General-purpose epilepsy ontologies are valuable for harmonizing seizure terminology and supporting informatics tools, but syndrome-focused applications increasingly require deeper representation of disease-specific concepts, especially when those concepts cross clinical and preclinical research settings.^9, 10^ Our prior work established the Epilepsy and Seizure Ontology (EpSO) as a formal epilepsy ontology for clinical research and patient care.9 However, the growth of DS-specific research, disease-modifying therapeutics,^11, 12^ genotype-phenotype interpretation, and translational model systems created the need for a more specialized semantic layer oriented to Dravet syndrome (DS).

### Objective

The objective of this work was to create a DS-focused ontology by extending EpSO into a computable representation of DS that could support standardized data capture, interoperable research assets, and downstream AI and analytics applications. We sought to preserve compatibility with the existing EpSO architecture while substantially broadening DS-specific coverage.

## Methods

The ontology engineering process began with the previously published epilepsy ontology as the parent semantic framework. That prior resource already provided reusable epilepsy-wide coverage for seizure terminology, syndromes, electrophysiology, anatomy, medications, and selected genetics, making it an appropriate substrate for syndrome-focused extension. Rather than create an isolated ontology, we used a modular extension strategy so that DS-specific content could inherit from, align with, and refine existing epilepsy concepts while preserving interoperability with prior EpSO-based tools and annotations. The domain content of the ontology was organized around nine expert-defined axes reflecting the major knowledge domains of Dravet syndrome: seizures, development, behavior, SUDEP, genetics, comorbidities, electrophysiology, pharmacology, and drug responsiveness; representative content for each domain is summarized in Table 1. These axes were established through iterative review by the scientific advisory board and were used to guide term expansion, hierarchical modeling, and prioritization of syndrome-specific content. Candidate additions were identified from expert knowledge, existing EpSO gaps, and recurring terms used in DS clinical and translational research workflows.

**Table 1.** Dravet syndrome ontology scope by domain and representative content.

| Axis | Scope in the Dravet syndrome ontology | Example concepts represented |
| --- | --- | --- |
| Seizures | Captures seizure types, triggers, temporal patterns, severity, and seizure-related clinical features relevant to Dravet syndrome | febrile seizure, hemiclonic seizure, myoclonic seizure, status epilepticus, temperature-induced seizure |
| Development | Represents neurodevelopmental trajectory and developmental impairments associated with Dravet syndrome | developmental delay, language impairment, cognitive impairment, developmental regression |
| Behavior | Encodes behavioral and neuropsychiatric manifestations observed in affected individuals | autistic features, hyperactivity, aggression, emotional dysregulation, attention difficulties |
| SUDEP | Represents concepts related to sudden unexpected death in epilepsy and associated risk-related knowledge | SUDEP, risk factors, seizure burden associations, nocturnal generalized seizures |
| Genetics | Organizes genes, variants, pathogenicity-related concepts, and genotype–phenotype relationships | SCN1A, pathogenic variant, sequence variant, gene attribute |
| Comorbidities | Captures associated neurologic, systemic, and functional conditions that co-occur with Dravet syndrome | sleep disturbance, gait abnormality, infections, autonomic dysfunction, gastrointestinal problems |
| Electrophysiology | Represents EEG patterns, epileptiform abnormalities, and related neurophysiologic findings | spike, sharp wave, polyspike, interictal discharge, photoparoxysmal response |
| Pharmacology | Encodes medications, treatment classes, and therapeutic interventions used in Dravet syndrome | valproate, clobazam, stiripentol, cannabidiol, fenfluramine |
| Drug responsiveness | Represents treatment response, resistance, efficacy, tolerability, and differential outcome patterns | drug-resistant epilepsy, partial response, seizure reduction, worsening, adverse effects |

Term curation proceeded domain by domain. For each proposed concept, the group assessed whether the content should be represented as a new class, modeled as a subclass of an existing epilepsy concept, or handled through synonym expansion or annotation. Review criteria included clinical relevance, semantic distinctness, expected reuse in data annotation, compatibility with existing hierarchy, and value for downstream applications such as registry harmonization, literature curation, knowledge graph construction, and AI-assisted retrieval. Duplicate or overly granular proposals were consolidated, while concepts judged essential for DS-specific representation were retained even when their use was expected to be concentrated in a single subcommunity such as basic science or genotype-phenotype studies.

The modeling strategy emphasized clinically meaningful and translationally useful constructs rather than a purely encyclopedic list of disease facts. Examples included temperature-sensitive seizure triggers and febrile events; developmental and age-stratified descriptors spanning neonate through adult; behavioral and neuropsychiatric features; respiratory, cardiac, and peri-ictal concepts relevant to premature mortality risk; gene-, variant-, and molecular consequence-level entities centered on SCN1A and related genes; treatment concepts including anti-seizure medications and treatment response patterns; and preclinical entities describing model systems, experimental conditions, and outcome measures. This allowed the ontology to capture how DS is studied and managed across clinics, laboratories, and informatics settings.

Ontology implementation followed OWL-based practices consistent with prior epilepsy ontology development and broader biomedical ontology design principles.^13-18^ Classes were added and organized in Protege; hierarchical placement and reuse decisions were reviewed iteratively; and ontology consistency was checked after each major round of edits. Baseline comparison used the legacy EpSO metrics panel available before DS-focused augmentation, which reported 7,916 axioms, 2,609 logical axioms, 1,961 classes, 45 object properties, 2 data properties, 2 individuals, and 7 annotation properties. The post-extension ontology was then compared with the public BioPortal release to document structural growth and confirm public dissemination of the updated artifact. Where relevant, the extension remained compatible with external resources already leveraged by EpSO, including anatomy and medication terminologies.^9, 19, 20^ After iterative review and refinement, the updated ontology was released through NCBO BioPortal as the current EpSO public version.

To evaluate the DS-focused extension, we used a multi-level assessment strategy. First, we performed structural assessment by comparing pre-extension EpSO metrics with the current public BioPortal release after DS-focused augmentation. Second, candidate terms and hierarchical placement were reviewed through an expert-guided curation process focused on clinical relevance, semantic distinctness, expected reuse, compatibility with the existing EpSO hierarchy, and appropriate granularity. Third, we assessed downstream task-based utility by reviewing informatics applications in which the ontology was used as an operational semantic resource. These included two published studies: an ontology-augmented large language model pipeline for extraction of drug-efficacy evidence from DS literature and an ontology-mediated large language model workflow for extraction of electrophysiologic findings across human and preclinical DS studies. We also considered the ontology’s role as the schema and semantic organization layer for our ongoing DS knowledge graph and AI assistant platform; because this work evaluates an ontology-derived knowledge graph and question-answering system rather than the ontology alone, we treated it as downstream implementation evidence rather than a standalone ontology benchmark.

For descriptive ontology profiling, the final RDF/OWL artifact was parsed to quantify named classes, local EpSO classes, object properties, datatype properties, annotation properties, subclass edges, top-level branches, hierarchy depth, leaf classes, and annotation counts. Top-level class composition was calculated by assigning each named local EpSO class to its highest local ancestor in the asserted subclass hierarchy. Because Dravet syndrome-relevant concepts were embedded across multiple EpSO branches rather than maintained as a formally tagged DS-only module, we report quantitative composition for the full ontology and describe the DS-focused extension as a syndrome-oriented semantic layer distributed across seizure, development, behavior, SUDEP/autonomic risk, genetics, comorbidity, electrophysiology, pharmacology, drug responsiveness, and model-system content.

## Results

The resulting ontology substantially broadened DS representation beyond the original epilepsy-centric baseline. Advisory board review converged on a syndrome-focused structure spanning nine major content domains: seizures, development, behavior, SUDEP/autonomic risk, genetics, comorbidities, electrophysiology, pharmacology, and drug responsiveness. These additions transformed the ontology from a general epilepsy resource with limited DS coverage into a more comprehensive semantic framework capable of representing the multidimensional phenotype and translational research ecosystem of DS. EpSO version 3.1 is a comprehensive, domain-specific ontology developed to represent epilepsy and related conditions, with a particular emphasis on Dravet syndrome (DS). The ontology comprises 14,050 RDF triples and 2,198 named URI classes, of which 2,189 are defined within the EpSO namespace. In addition to these named classes, the ontology incorporates 294 class expressions or logical restrictions, reflecting the use of OWL constructs to encode structured relationships and constraints. EpSO includes 53 object properties, 5 datatype properties, and 5 annotation properties. Of the object properties, 51 are defined within EpSO, indicating that most relationships used for domain modeling were specifically designed to capture epilepsy-relevant semantics. The ontology further includes 2,159 subclass relationships, 2,149 rdfs:label annotations, 681 rdfs:comment annotations, 1,415 rdfs:seeAlso annotations, and 111 synonym annotations. Overall ontology growth relative to the legacy EpSO baseline is summarized in Table 2.

**Table 2.** Selected ontology growth indicators before and after Dravet syndrome-focused extension.

| Indicator | Legacy EpSO baseline | Current public release | Interpretation |
| --- | --- | --- | --- |
| Classes | 1,961 | 2,198 | Broader concept coverage after DS-focused expansion |
| Properties | 45 object properties; 2 data properties; 7 annotation properties | 59 BioPortal-indexed properties; direct RDF parsing identified 53 object, 5 datatype, and 5 annotation properties | Expanded relation and annotation structure for DS-focused modeling |
| Individuals | 2 | 2 | No meaningful change in individuals count |
| Release status | Published EpSO foundation | BioPortal EPSO v3.1 (2025) | Publicly released and discoverable ontology artifact |
| Domain emphasis | General epilepsy and seizure informatics | Expanded DS clinical + preclinical content | Specialization supports syndrome-specific translational work |

The class hierarchy is organized into 30 top-level branches that collectively define the conceptual scope of the ontology. The largest branches include Bodily Feature, Clinical Drug Component, Physiological Process, Disorder, Etiology, Gene, EEG Pattern, Electrode, Biological Substance, Physiological Data, Genomic Variation, Model System, Measurement Method, and Measurement Unit. These branches demonstrate that EpSO extends beyond traditional epilepsy classifications to encompass a broad semantic framework integrating clinical phenotypes, anatomical and physiological structures, electrophysiological patterns, pharmacological entities, genetic and genomic information, etiological factors, measurement concepts, and experimental systems. The Bodily Feature branch contains 532 classes, reflecting extensive coverage of anatomical and phenotypic characteristics relevant to epilepsy.

Clinical Drug Component includes 320 classes representing drug ingredients, formulations, and medication-related concepts. Physiological process contains 233 classes, while Disorder and Etiology include 172 and 168 classes, respectively. Gene and Genomic Variation branches contain 155 and 48 classes, supporting representation of gene-level and variant-level knowledge. Figure 1 illustrates this hierarchical class composition, showing the distribution of named local EpSO classes across major top-level branches and selected second-level sub-branches.

**Figure 1.**
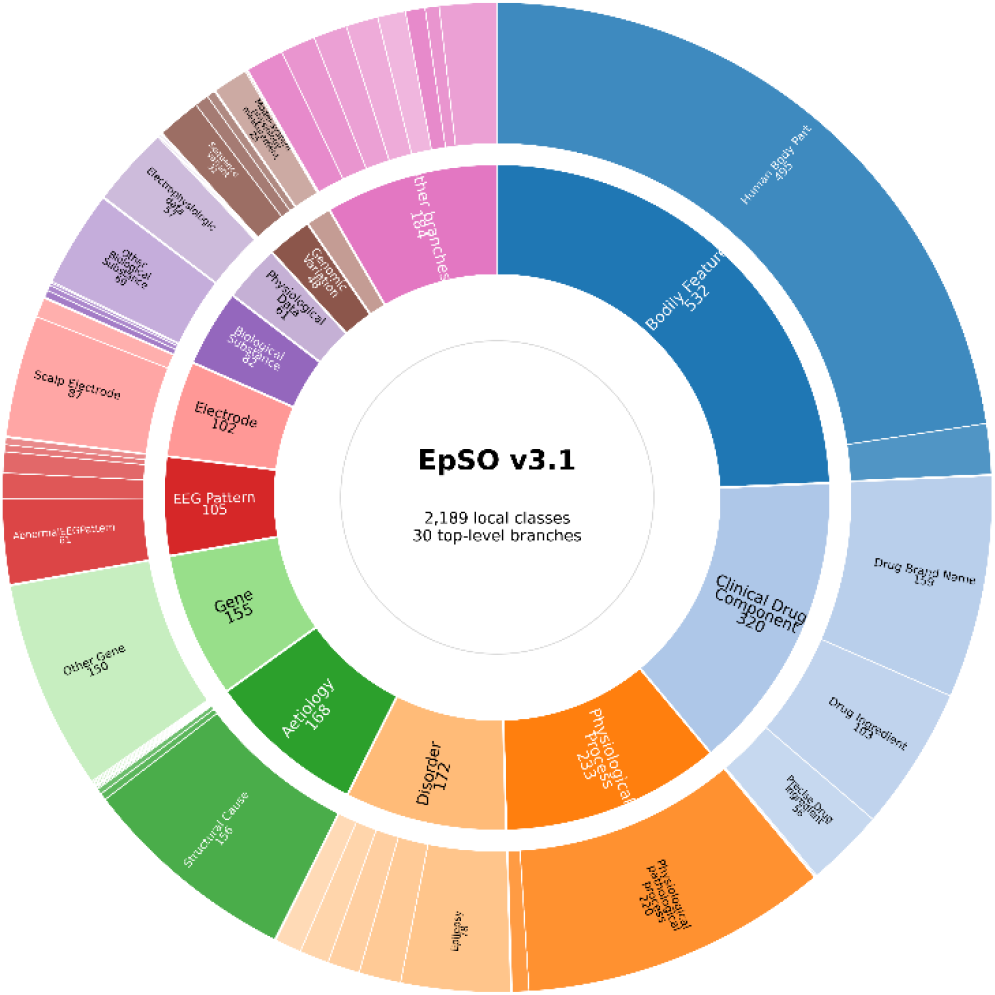
Hierarchical class composition of EpSO 3.1. The sunburst plot summarizes the distribution of named local EpSO classes across major top-level branches and selected second-level sub-branches. Segment size is proportional to class count. The figure highlights EpSO’s multidomain structure, spanning clinical phenotype, medications, physiologic processes, disorders, etiologic concepts, genetics, electrophysiology, genomic variation, and measurement concepts. DS-focused content is embedded across multiple ontology branches rather than represented as a single isolated subtree.

Within this structure, the Dravet syndrome-focused extension is not represented as a single isolated branch. Instead, Dravet syndrome is modeled as a disease class within the broader epilepsy disorder hierarchy, while DS-relevant concepts are distributed across multiple branches of the ontology. This design reflects the multifaceted nature of DS, which encompasses seizure phenotypes, environmental triggers such as temperature sensitivity, developmental and behavioral features, autonomic dysfunction and SUDEP risk, genetic and genomic variation, electrophysiological findings, pharmacological treatments, treatment response patterns, and experimental model systems. By embedding DS-related concepts throughout the ontology, EpSO enables integrated representation of syndrome-specific knowledge while maintaining interoperability with general epilepsy, neurophysiology, genetics, pharmacology, and experimental research domains; this distributed DS-focused semantic layer is summarized in Figure 2.

**Figure 2.**
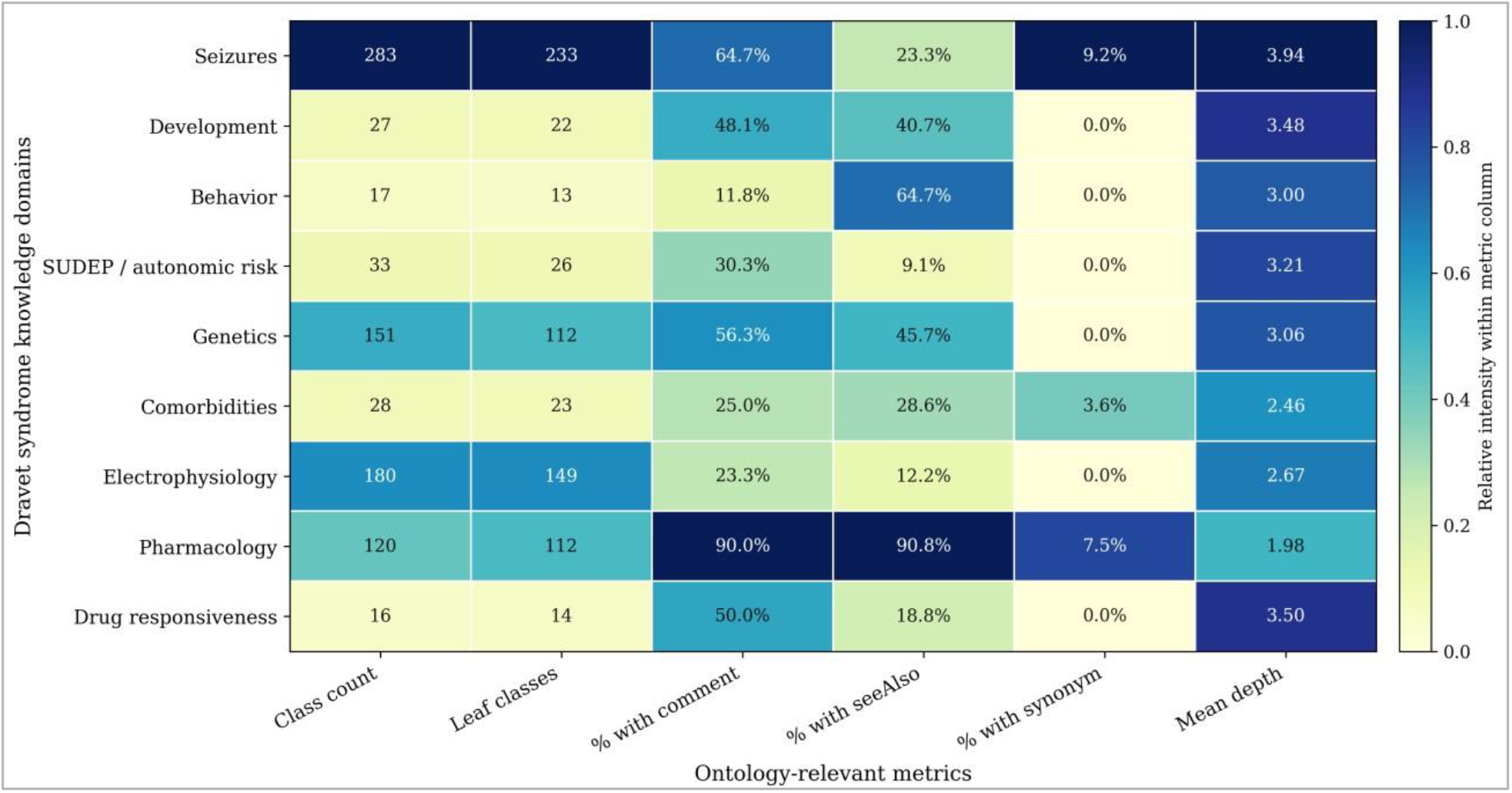
Dravet syndrome semantic coverage across EpSO v3.1 knowledge domains. The heatmap summarizes Dravet syndrome-relevant ontology content across nine expert-defined domains: seizures, development, behavior, SUDEP/autonomic risk, genetics, comorbidities, electrophysiology, pharmacology, and drug responsiveness. Rows correspond to DS knowledge domains and columns report ontology-relevant metrics, including class count, number of leaf classes, percentage of classes with comments, percentage with rdfs:seeAlso annotations, percentage with synonym annotations, and mean hierarchy depth. A total of 715 unique local EpSO classes were matched to one or more DS-relevant domains using expert-curated keyword groupings; because DS concepts are embedded across the ontology rather than maintained as a single isolated subtree, a class could contribute to more than one domain. Color intensity is normalized within each metric column. This figure demonstrates that the DS-focused extension functions as a distributed semantic layer spanning clinical phenotype, genetics, electrophysiology, pharmacology, treatment response, SUDEP/autonomic risk, and related translational content.

The ontology exhibits a broad yet structured hierarchy, with 30 top-level branches, 1,761 leaf classes, a mean class depth of 4.46, and a maximum depth of 16. This organization balances granularity with navigability, enabling both detailed representation and intuitive exploration. The large number of leaf classes reflects a high degree of specificity, while the distribution across multiple top-level branches supports access through clinically meaningful categories such as disorders, physiological features, genes, medications, EEG patterns, and model systems. Annotation properties, including labels, comments, cross-references, and synonyms, enhance interpretability, traceability, and usability for both human users and computational applications. Class-level annotation completeness is summarized in Figure 3.

**Figure 3.**
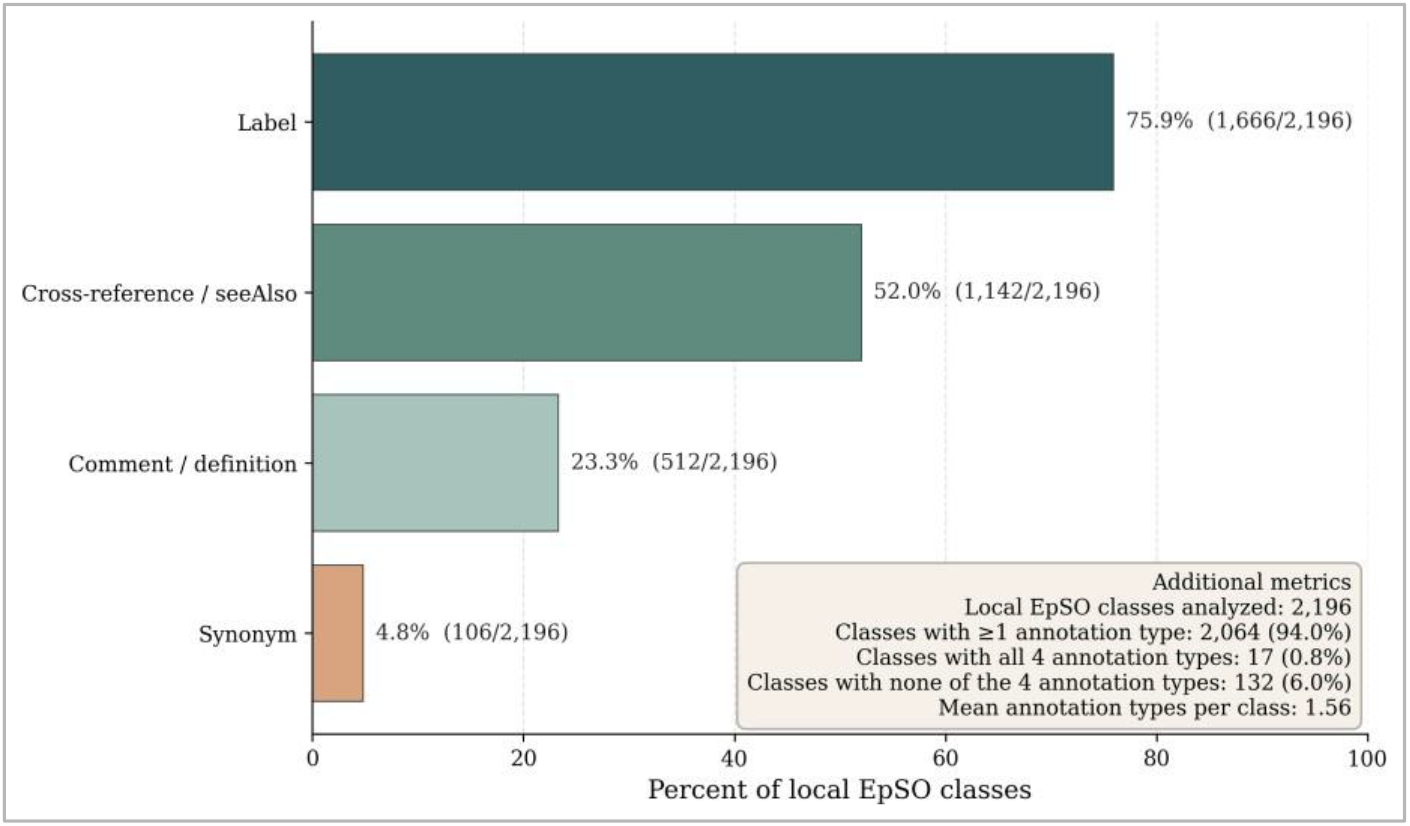
Annotation completeness profile of EpSO v3.1. The bar chart summarizes class-level annotation completeness across 2,196 local EpSO classes in the RDF/OWL ontology artifact. Metrics include the proportion of classes with an rdfs:label, rdfs:comment or definition-like annotation, rdfs:seeAlso or cross-reference annotation, and synonym annotation. Overall, 2,064 classes (94.0%) contained at least one of the four annotation types, whereas 132 classes (6.0%) contained none of these annotations. These metrics provide a curation-quality profile of EpSO v3.1 and characterize the ontology’s readiness for human interpretation, semantic review, data annotation, literature mining, and ontology-grounded AI workflows.

This quantitative representation provides a clear overview of the ontology’s structure and content. The distribution of classes across branches highlights the ontology’s multidomain design, with substantial representation in bodily and phenotypic features, medication-related entities, physiological processes, disorders, etiological concepts, genes, EEG patterns, and experimental or measurement-related domains. This structure aligns with the goal of extending EpSO into a comprehensive semantic resource capable of supporting DS-focused clinical, translational, and artificial intelligence applications.

The largest qualitative gains were observed in domains that are typically underrepresented in generic epilepsy schemas. Developmental representation was expanded to better capture age-linked clinical course and neurologic assessment; behavioral content was broadened to include neuropsychiatric and school-function descriptors; genetics was deepened from general epilepsy-genecoverage toward DS-relevant genes, variants, and molecular consequences; and SUDEP-related content was made more granular by incorporating respiratory, cardiac, postural, and peri-ictal concepts discussed in DS research. The extension also added explicit preclinical vocabulary,enabling representation of murine and other experimental DS studies alongside human clinical knowledge.

From an informatics perspective, the DS-focused extension improved the ontology’s utility in three ways: it provided a more appropriate schema for DS-specific structured data capture and cross-study harmonization; it supplied a controlled vocabulary for annotating literature, datasets, and experimental findings; and it created a domain structure that could be reused in knowledge graphs, retrieval-augmented generation, and ontology-grounded AI workflows. Because the DS-focused content was incorporated through extension rather than replacement, these gains were achieved while preserving continuity with the broader epilepsy ontology framework. Task-based reuse provided additional evidence that the ontology was usable beyond structural expansion. In a published drug-efficacy extraction study, the ontology was embedded into a phased in-context learning workflow for Gemini-based extraction of DS treatment evidence. The ontology supplied structured terms for anti-seizure medications, seizure triggers, model systems, and treatment-response concepts; ontology-augmented prompting reproduced a benchmark dataset for 17 anti-seizure medications with 100% accuracy and was subsequently scaled to 4,935 full-text DS articles. This represented a task-based validation of the ontology as a semantic scaffold for literature-scale extraction and translational evidence synthesis.^21^

In a second published study, the electrophysiology portion of the ontology was used to guide large language model extraction of EEG and seizure-related findings from a curated corpus of 47 DS articles spanning human, zebrafish, and mouse studies. In this application, ontology-mediated prompting organized terms for organisms, seizure trigger types, ictal and interictal findings, and EEG-pattern concepts. This use case provided domain-specific evidence that the ontology could harmonize electrophysiologic findings across clinical and preclinical literature.^22^ The same ontology is also being used as the schema and semantic organization layer for our ongoing DS knowledge graph and AI assistant platform. In that project, ontology classes and relationships guide knowledge graph construction, retrieval, and contextual grounding of AI-generated responses. We have developed a benchmark for evaluating this ontology-derived knowledge graph and question-answering workflow; however, because that benchmark evaluates the combined knowledge graph and AI retrieval system rather than the ontology in isolation, we interpret it as downstream implementation evidence rather than as a formal ontology-only validation. Together, these structural, expert-guided, published task-based, and ongoing implementation assessments provide complementary evidence of ontology evaluation and utility, as summarized in Table 3.

**Table 3.** Summary of structural, expert-guided, and task-based evidence supporting DS ontology evaluation.

| Evidence type | Source or process | Ontology role | Quantitative/systematic element | Interpretation |
| --- | --- | --- | --- | --- |
| Structural assessment | Legacy EpSO vs BioPortal EPSO v3.1 | DS-focused extension of parent epilepsy ontology | Classes increased from 1,961 to 2,198; properties increased from 45 object properties to 59 properties | Demonstrates measurable representational growth |
| Expert-guided curation | Scientific advisory board and iterative review | Term selection, hierarchy, granularity, and domain scope | Domain-by-domain review across nine DS axes | Supports clinical and translational relevance |
| Published task-based reuse | Drug-efficacy LLM extraction study | Prompt scaffold and controlled vocabulary for DS treatment evidence | 17 ASM benchmark reproduced with 100% accuracy; scaled to 4,935 full-text articles | Demonstrates utility for literature-scale extraction |
| Published domain-specific reuse | Electrophysiology LLM extraction study | EEG, seizure-trigger, organism, and model-system organization | Curated corpus of 47 DS electrophysiology articles | Demonstrates harmonization across human and preclinical evidence |
| Ongoing downstream implementation | DS-KG/KroMA platform | Ontology-derived KG schema and retrieval layer | Benchmark developed for KG/QA evaluation | Supports implementation value, but not ontology-only validation |

The final result is therefore not simply a larger ontology, but a more specialized one. The added content improves coverage of syndrome-specific concepts that clinicians, translational investigators, and data scientists routinely need but that were only partially represented in the original resource. This specialization is important for diseases such as DS, where meaningful interpretation depends on relating seizures to genotype, development, behavior, treatment response, autonomic vulnerability, and preclinical evidence within a single computable semantic model.

## Discussion

This work addresses an important gap between broad epilepsy ontologies and the specialized semantic requirements of a single high-impact syndrome. Dravet syndrome is more than a seizure disorder; it is a longitudinal, multisystem condition in which meaningful data arise from genetics, neurodevelopment, electrophysiology, therapeutics, adverse effects, autonomic dysfunction, and mortality-related mechanisms.^1, 2^ A syndrome-specific ontology is therefore useful not only for terminology standardization, but also for structuring the knowledge interfaces among clinicians, basic scientists, data analysts, and AI systems.

The decision to extend EpSO rather than build an unrelated standalone ontology preserved interoperability with prior epilepsy informatics work and reduced duplication of already-curated seizure, anatomy, medication, electrophysiology, and genetics concepts.^9, 19, 20^ This strategy is aligned with modular biomedical ontology development, in which specialized disease content is added through principled reuse of existing artifacts rather than creating disconnected vocabularies.^14^ For a condition such as DS, this is particularly important because the syndrome intersects with multiple biomedical domains. DS cannot be represented adequately by a single disease node or a narrow diagnostic hierarchy. Instead, its representation requires coordination across seizure types, seizure triggers, developmental and behavioral features, SUDEP and autonomic risk, genes and variants, EEG findings, drug exposure, treatment response, and preclinical model systems.

The RDF-based descriptive analysis further supports this interpretation. EpSO version 3.1 is not simply a disease list or seizure classification resource; it is a multidomain semantic framework spanning bodily and phenotypic features, medication-related entities, physiologic processes, disorders, etiologic concepts, genes, EEG patterns, biological substances, physiologic data, genomic variation, model systems, and measurement concepts. The largest ontology branches reflect the domains needed to represent both clinical and translational epilepsy knowledge. This structure is well suited to DS research, where clinical observations, molecular mechanisms, electrophysiologic patterns, therapeutic evidence, and experimental model findings often need to be interpreted together. More broadly, this reflects the role of biomedical ontologies in enabling structured, computable representation of heterogeneous health-system and translational data.^23-25^ The ontology therefore provides a semantic bridge between clinical and preclinical DS knowledge rather than functioning only as a controlled vocabulary for syndrome labels.

An important implication of this structure is that the Dravet syndrome-focused extension should be understood as an embedded semantic layer within EpSO rather than as a separate Dravet-only subtree. In the ontology artifact, Dravet syndrome is represented as a disease class within the broader epilepsy disorder hierarchy, while DS-relevant content is distributed across multiple branches. This modeling choice supports interoperability because DS-related concepts remain connected to broader epilepsy, genetics, pharmacology, physiology, and model-system structures. It also better reflects the real-world organization of DS knowledge, where the syndrome is defined not only by diagnosis but also by characteristic seizure phenotypes, developmental trajectory, genetic etiology, treatment response, and translational research models.

The DS-focused extension also has immediate practical relevance. It can underpin DS registries, support article and dataset annotation, organize preclinical findings, and provide a schema for DS knowledge graphs and LLM-based retrieval or reasoning systems.^5, 24, 25^ Because the ontology explicitly models drug responsiveness and adverse effects, it may also improve the comparability of therapeutic evidence across clinical studies and laboratory models.^11, 12^ This is especially important for rare diseases, where evidence is often distributed across small clinical cohorts, case series, preclinical studies, and emerging therapeutic reports.

Importantly, the ontology has already moved into real DS applications. Its reuse in published drug-efficacy and electrophysiology studies provides early evidence that the ontology is sufficiently mature to support literature-scale NLP and LLM workflows, while its ongoing use in our DS knowledge graph and AI assistant platform suggests a path toward interactive DS question answering and knowledge navigation.^21, 22^ Together, these use cases strengthen the argument that ontology development for rare disease should be evaluated not only by counts of classes and properties, but also by the diversity of downstream tasks it can support.

Several limitations should be noted. Although this study includes structural comparison, RDF-based descriptive profiling, expert-guided curation, and downstream task-based reuse, it does not yet include a dedicated ontology-only benchmark based on independently specified competency questions, inter-annotator agreement, systematic cross-ontology mapping, or formal coverage testing against an external DS corpus.^17, 18^ The published downstream studies and ongoing DS knowledge graph work support the ontology’s practical utility, but they evaluate ontology-enabled workflows rather than the ontology as an isolated artifact. In addition, because DS-relevant classes are embedded across multiple EpSO branches rather than tagged as members of a formal Dravet-specific module, the current artifact does not support a precise automated count of Dravet-only classes without additional curation metadata. This is not necessarily a modeling weakness, because embedding DS concepts within shared epilepsy, genetics, pharmacology, electrophysiology, and model-system hierarchies promotes interoperability. However, future releases would benefit from explicit module-level annotations or subset tags identifying DS-focused additions. Such tags would support versioned growth tracking, disease-specific coverage analysis, governance, and automated reporting of module-level ontology statistics.

Future work will focus on formal coverage evaluation, continued versioned governance as DS knowledge evolves, and assessment of whether this specialization workflow can be scaled to other rare diseases requiring integration of clinical, genetic, therapeutic, and translational knowledge. Additional evaluation should include competency-question testing, expert annotation studies, assessment of inter-annotator agreement, and comparison with related biomedical ontologies where relevant.^17, 18^ Public ontology dissemination through resources such as BioPortal can improve discoverability and reuse,^24, 26^ while standardized ontology engineering and release workflows, including tools such as Protege, ROBOT, and the Ontology Development Kit, can support more transparent quality control, versioning, and long-term maintenance.^15, 17, 18, 24-30^ These steps would strengthen EpSO as a sustainable rare disease informatics resource and would further clarify how disease-focused ontology specialization can support AI-enabled biomedical knowledge systems.

## Conclusion

We developed a Dravet syndrome ontology through expert-guided domain specialization and organized the resulting resource around nine clinically and translationally important DS domains. By extending an established epilepsy ontology rather than replacing it, the work preserved interoperability while substantially improving representation of DS-specific concepts across clinical care, translational research, preclinical studies, and AI-enabled knowledge applications. The resulting ontology provides a practical semantic foundation for DS data harmonization and future DS informatics infrastructure.

## Data Availability

All data produced are available online at https://bioportal.bioontology.org/ontologies/EPSO

## Acknowledgements

This work was supported in part by the U.S. National Institutes of Health (NIH) under grants U24EB029005 and R01DA053028; the U.S. Department of Defense (DoD) under grant W81XWH2110859; the Clinical and Translational Science Collaborative of Cleveland, funded by the NIH National Center for Advancing Translational Sciences through Clinical and Translational Science Award UL1TR002548; and the CURE SUDEP grant.

## References

1. Wirrell EC, Hood V, Knupp KG, et al. International consensus on diagnosis and management of Dravet syndrome. Epilepsia 2022;63(7):1761–77 doi: 10.1111/epi.17274

2. Miller IO, Sotero de Menezes MA. SCN1A seizure disorders. 2022

3. Dravet C. The core Dravet syndrome phenotype. Epilepsia 2011;52 Suppl 2:3–9 doi: 10.1111/j.1528-1167.2011.02994.x

4. Claes L, Del-Favero J, Ceulemans B, et al. De novo mutations in the sodium-channel gene SCN1A cause severe myoclonic epilepsy of infancy. Am J Hum Genet 2001;68(6):1327–32 doi: 10.1086/320609

5. Strzelczyk A, Lagae L, Wilmshurst JM, et al. Dravet syndrome: A systematic literature review of the illness burden. Epilepsia Open 2023;8(4):1256–70 doi: 10.1002/epi4.12832

6. Villas N, Meskis MA, Goodliffe S. Dravet syndrome: Characteristics, comorbidities, and caregiver concerns. Epilepsy Behav 2017;74:81–86 doi: 10.1016/j.yebeh.2017.06.031

7. Licheni SH, McMahon JM, Schneider AL, et al. Sleep problems in Dravet syndrome: a modifiable comorbidity. Dev Med Child Neurol 2018;60(2):192–98 doi: 10.1111/dmcn.13601

8. Kearney J. Sudden unexpected death in dravet syndrome. Epilepsy Curr 2013;13(6):264–5 doi: 10.5698/1535-7597-13.6.264

9. Sahoo SS, Lhatoo SD, Gupta DK, et al. Epilepsy and seizure ontology: towards an epilepsy informatics infrastructure for clinical research and patient care. J Am Med Inform Assoc 2014;21(1):82–9 doi: 10.1136/amiajnl-2013-001696

10. Scheffer IE, Berkovic S, Capovilla G, et al. ILAE classification of the epilepsies: Position paper of the ILAE Commission for Classification and Terminology. Epilepsia 2017;58(4):512–21 doi: 10.1111/epi.13709

11. Wirrell EC, Nabbout R. Recent Advances in the Drug Treatment of Dravet Syndrome. CNS Drugs 2019;33(9):867–81 doi: 10.1007/s40263-019-00666-8

12. Devinsky O, Cross JH, Laux L, et al. Trial of Cannabidiol for Drug-Resistant Seizures in the Dravet Syndrome. N Engl J Med 2017;376(21):2011–20 doi: 10.1056/NEJMoa1611618

13. Hitzler P, Krötzsch M, Parsia B, et al. OWL 2 web ontology language primer. W3C recommendation 2009;27(1):123

14. Smith B, Ashburner M, Rosse C, et al. The OBO Foundry: coordinated evolution of ontologies to support biomedical data integration. Nat Biotechnol 2007;25(11):1251–5 doi: 10.1038/nbt1346

15. Gruber TR. A translation approach to portable ontology specifications. Knowledge acquisition 1993;5(2):199–220

16. Noy NF, McGuinness DL. Ontology Development 101: A Guide to Creating Your First Ontology. Stanford Knowledge Systems Laboratory Technical Report KSL-01-05 and Stanford Medical Informatics Technical Report SMI-2001-0880. 2001.

17. Grüninger M, Fox MS. Methodology for the Design and Evaluation of Ontologies. In: Proceedings of the Workshop on Basic Ontological Issues in Knowledge Sharing, IJCAI; 1995

18. Amith M, He Z, Bian J, et al. Assessing the practice of biomedical ontology evaluation: Gaps and opportunities. J Biomed Inform 2018;80:1–13 doi: 10.1016/j.jbi.2018.02.010

19. Rosse C, Mejino JL, Jr. A reference ontology for biomedical informatics: the Foundational Model of Anatomy. J Biomed Inform 2003;36(6):478–500 doi: 10.1016/j.jbi.2003.11.007

20. Nelson SJ, Zeng K, Kilbourne J, et al. Normalized names for clinical drugs: RxNorm at 6 years. J Am Med Inform Assoc 2011;18(4):441–8 doi: 10.1136/amiajnl-2011-000116

21. Golnari P, Prantzalos K, Hood V, et al. Ontology accelerates few-shot learning capability of large language model: A study in extraction of drug efficacy in a rare pediatric epilepsy. Int J Med Inform 2025;201:105942 doi: 10.1016/j.ijmedinf.2025.105942

22. Golnari P, Prantzalos K, Upadhyaya D, et al. Human in the Loop: Embedding Medical Expert Input in Large Language Models for Clinical Applications. Stud Health Technol Inform 2025;329:658–62 doi:10.3233/SHTI250922

23. Callahan TJ, Stefanski AL, Wyrwa JM, et al. Ontologizing health systems data at scale: making translational discovery a reality. NPJ Digit Med 2023;6(1):89 doi: 10.1038/s41746-023-00830-x

24. Wilkinson MD, Dumontier M, Aalbersberg IJ, et al. The FAIR Guiding Principles for scientific data management and stewardship. Scientific data 2016;3(1):1–9

25. Bodenreider O. Biomedical ontologies in action: role in knowledge management, data integration and decision support. Yearbook of medical informatics 2008;17(01):67–79

26. Whetzel PL, Noy NF, Shah NH, et al. BioPortal: enhanced functionality via new Web services from the National Center for Biomedical Ontology to access and use ontologies in software applications. Nucleic Acids Res 2011;39(Web Server issue):W541–5 doi: 10.1093/nar/gkr469

27. Jackson RC, Balhoff JP, Douglass E, et al. ROBOT: A Tool for Automating Ontology Workflows. BMC Bioinformatics 2019;20(1):407 doi: 10.1186/s12859-019-3002-3

28. Matentzoglu N, Goutte-Gattat D, Tan SZK, et al. Ontology Development Kit: a toolkit for building, maintaining and standardizing biomedical ontologies. Database 2022;2022:baac087

29. Musen MA, Protege T. The Protege Project: A Look Back and a Look Forward. AI Matters 2015;1(4):4–12 doi: 10.1145/2757001.2757003

30. Noy N, Gao Y, Jain A, et al. Industry-scale Knowledge Graphs: Lessons and Challenges: Five diverse technology companies show how it’s done. Queue 2019;17(2):48–75.

